# Statistical Analysis Plan (SAP) – DREAM: an adaptive, randomised, placebo-controlled trial of duloxetine for reducing leg pain in people with chronic sciatica

**DOI:** 10.64898/2026.07.12.26357883

**Authors:** Xiaoqiu Liu, Laurent Billot, Anthony Devaux, Christopher Maher, Christine Lin, Richard O Day, Rowena Ivers, Martin Underwood, Andrew J McLachlan, Bethan Richards, Nanna B Finnerup, Cecilia Taing, Kate Tong, Masoud Jamshidi, Maliha Hassan, Melanie Hamilton, Emily Atkins, Giovanni Ferreira

## Abstract

DREAM is a randomised, superiority, parallel-group, placebo-controlled, participant, clinician, and assessor blinded trial with an adaptive group sequential design that allows early stopping for efficacy or futility. The purpose is to investigate whether taking 60 mg of duloxetine daily for 12 weeks in addition to guideline-recommended advice, compared with placebo in addition to guideline-recommended advice, can reduce leg pain intensity in individuals with chronic sciatica.

The primary outcome is leg pain intensity measured on a 0-10 numerical pain rating scale. It will be analysed using a repeated-measures linear mixed model. This statistical analysis plan pre-specifies the methods of analysis to be used in the interim analysis and the final analysis for the outcomes and key variables collected in the trial. It includes planned sensitivity analyses for the final analysis, including covariate adjustments and subgroup analyses, as well as the health economics analysis plan.

## 4 Background

Antidepressants are commonly used for chronic pain, including chronic sciatica (a commonly used term to describe painful radiculopathy) [2, 3]. However, the evidence of effectiveness in this population is inconclusive. A 2021 systematic review [4] found evidence that antidepressants such as duloxetine, a serotonin-norepinephrine reuptake inhibitor (SNRI), may be effective for chronic sciatica and provide clinically important pain reduction in the short-term. However, effect estimates were based on three small (n=96), low-quality trials, and the certainty of evidence ranged from low to very low. We are conducting the *Duloxetine for the treatment of chronic sciatica (DREAM)* trial to investigate the effectiveness and safety of duloxetine, compared to placebo, in people with chronic sciatica [1]. Here we describe the statistical analysis plan that has been developed and signed off by DREAM investigators before the first pre-specified interim analysis.

### 4.1 Study synopsis

DREAM trial is an investigator-initiated randomised, parallel-group superiority, placebo-controlled, participant, clinician, and assessor blinded trial with an adaptive group sequential design investigating the efficacy and safety of duloxetine in participants with sciatica of at least 3 months duration. Participants are randomised at a 1:1 ratio to duloxetine or placebo. It is planned to recruit 332 participants from general practices, specialist clinics, hospital emergency departments, hospital in-patient wards, and from the community. In the active treatment group, participants receive duloxetine 60 mg per day for 12 weeks, including 1 week of titration at 30 mg/day. The treatment phase is followed by a 2-week tapering phase where they receive duloxetine 30 mg/day. In the placebo group, participants receive identical placebo tablets titrated to the same regimen. Participants in both groups will receive guideline-recommended advice from their study doctor, consisting of information on the nature of sciatica, its prognosis, encouragement to continue with normal activities, importance of avoiding bed rest, and to avoid opioid medicines.

Participants are followed up for 52 weeks, with outcomes measured at 4, 8, 12, 16, 26, and 52 weeks post-randomisation. The primary outcome is leg pain intensity. The primary time point is 12 weeks post-randomisation. Secondary outcomes include back pain intensity, disability, time to recovery, quality of life, depressive and anxiety symptoms, work absenteeism, and sleep disturbance. Adverse events are recorded, and a cost-effectiveness analysis will be conducted.

### 4.2 Ethics, sponsorship, and trial registration

Approval was granted by The University of Sydney Human Research Ethics Committee (2024/HE000160) and Sydney Local Health District Ethics Committee (2024/ETH02476) and registered prospectively with the Australian New Zealand Clinical Trials Registry (ACTRN12624000919516). The University of Sydney is the sponsor.

### 4.3 Study population

#### 4.3.1 Inclusion criteria

1. Adults (≥18 years) with radiating pain into one leg in a dermatomal distribution.
2. Leg pain duration of at least three months.
3. Evidence of nerve root involvement, defined by the presence of at least ONE of the following clinical signs in the corresponding distribution: myotomal weakness and/or diminished reflex and/or sensory deficit and/or imaging evidence of nerve root impingement that is consistent with the clinical presentation.
4. Leg pain that is at least moderate in intensity at the time of enrolment by a study doctor (as measured by a modified version of item 21 in the 36-Item Short Form Survey Instrument).
5. An adequate understanding of English or the availability of interpretation services for the participant to complete the trial.

#### 4.3.2 Exclusion criteria

1. Known or suspected specific pathologies in the spine (e.g. fracture, cauda equina syndrome).
2. Known or suspected malignancy.
3. Having had spinal surgery or other interventional procedure (e.g. epidural injection) in the preceding 6 months.
4. Scheduled to have a spinal procedure (e.g. spinal surgery, epidural injection) within 12 weeks at the time of enrolment.
5. Currently taking (or taken in last 2 weeks) any antidepressant for any condition.
6. Any contraindications to duloxetine: known hypersensitivity to duloxetine, known acute or chronic liver disease, concomitant use of CYP1A2 inhibitors (e.g. fluvoxamine) or concomitant use of monoamine oxidase inhibitors (e.g. moclobemide), or use that has ceased less than 2 weeks ago.
7. Known history of chronic kidney disease stage 4 or above.
8. Precautions for use of duloxetine as specified by the Product Information Sheet where risks outweigh potential benefits (e.g. depressive symptoms for which treatment is required as judged by the study doctor, bipolar disorder, history of seizure disorder, etc.).
9. Previous severe adverse reaction to duloxetine (e.g. serotonin syndrome) as judged by the study doctor.
10. For females: pregnant, breastfeeding, or planning conception during the treatment period.

### 4.4 Interventions

#### Intervention group

Participants randomised to the active arm of the study receive a 12-week course of oral duloxetine followed by a 2-week tapering phase before cessation. The starting dose is 30 mg/day for 1 week (one 30 mg capsule per day), increasing to 60 mg/day for 11 weeks (maintenance phase – two 30 mg capsules per day). In the 2-week tapering phase, they receive 30 mg/day (one 30 mg capsule per day) for 2 weeks before treatment is discontinued. Participants in this group also receive guideline-recommended advice as described in the trial protocol and other usual care as judged appropriate by their study doctor.[1] enrolling them into the study and prescribing the study treatment.

#### Control group

Participants randomised to the placebo arm of the study receive the equivalent number of matching placebo capsules throughout the 14 weeks of the treatment phase. Participants in this group also receive guideline-recommended advice as described in the trial protocol and other usual care as deemed appropriate by the study doctor. enrolling them into the study and prescribing the study treatment.

### 4.5 Study visits

**Figure 1.**
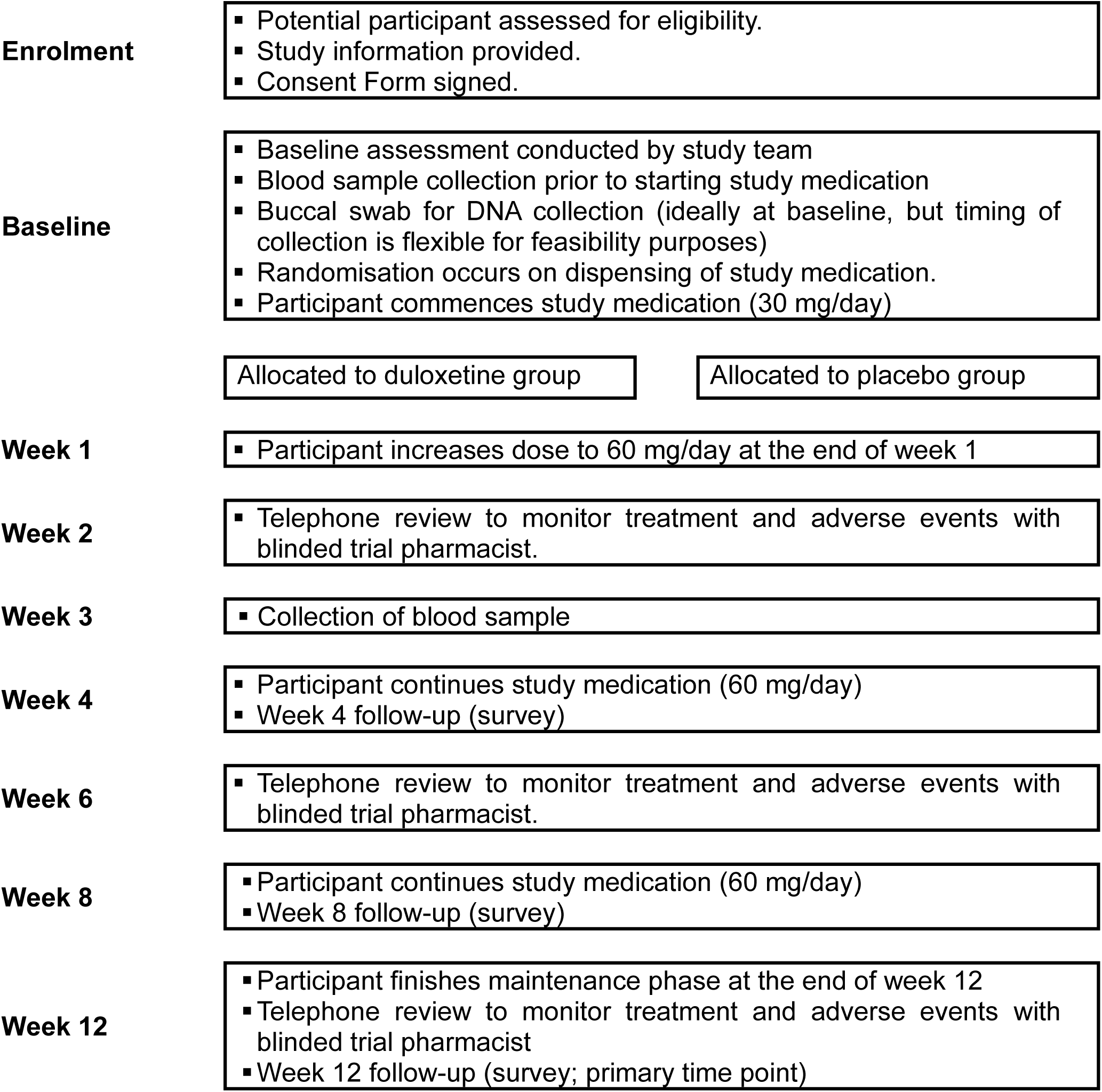
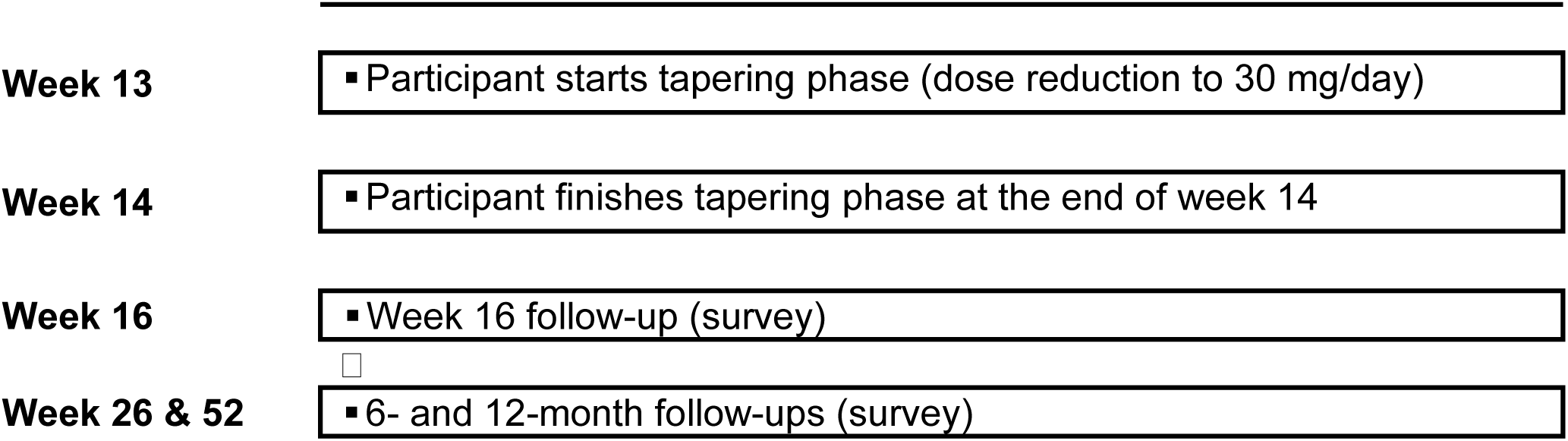
Study Flowchart.

**Table 1.** Study Visits.

| Interventions | Baseline | W1 | W2 | W3 | W4 | W6 | W8 | W12 | W13 | W14 | W16 | W26 | W52 |
| --- | --- | --- | --- | --- | --- | --- | --- | --- | --- | --- | --- | --- | --- |
| Screening | x |  |  |  |  |  |  |  |  |  |  |  |  |
| Eligibility | x |  |  |  |  |  |  |  |  |  |  |  |  |
| Informed Consent | x |  |  |  |  |  |  |  |  |  |  |  |  |
| Surveys | x |  |  |  | x |  | x | x |  |  | x | x | x |
| Treatment monitoring |  |  | x |  |  | x |  | x |  |  |  |  |  |
| AE/SAE assessment |  |  | x |  | x | x | x | x |  |  | x |  |  |
| Blood collection | x |  |  | x |  |  |  |  |  |  |  |  |  |
| Buccal DNA collection | x |  |  |  |  |  |  |  |  |  |  |  |  |
| Start medication |  | x |  |  |  |  |  |  |  |  |  |  |  |
| Dose escalation |  |  | x |  |  |  |  |  |  |  |  |  |  |
| Dose maintenance |  |  |  | x | x | x | x | x |  |  |  |  |  |
| Dose reduction |  |  |  |  |  |  |  |  | x | x |  |  |  |
| End of treatment |  |  |  |  |  |  |  |  |  | x |  |  |  |
| Pain and medication diary |  | Weeks 1-14 |  |  |  |  |  |  |  |  |  |  |  |

### 4.6 Outcomes

**Table 2.** Outcome and measurements.

**4.6 Outcomes****Table 2. Outcome and measurements**
| Outcome | Measurement tool | Measurement occasions |
| --- | --- | --- |
| **Primary outcome** |  |  |
| Leg pain intensity | Numerical pain rating scale (NPRS) (0-10, best to worst)<br>*“I would like you to rate your leg pain on a scale from 0 to 10 where 0 is no pain and 10 is the worst possible pain. Please give a number to describe your average pain over the last 24 hours.”* | Baseline, weeks 4, 8, 12, 16, 26, and 52<br>The primary outcome time point is 12 weeks post-randomisation |
| **Secondary outcomes** |  |  |
| Disability (key secondary outcome) | Roland Morris Disability Scale [5] for Sciatica (0-23, best to worst) | Baseline, weeks 4, 8, 12, 16, 26, and 52 |
| Low back pain intensity | Numerical pain rating scale (NPRS) (0-10, best to worst)<br>*“I would like you to rate your back pain on a scale from 0 to 10 where 0 is no pain and 10 is the worst possible pain. Please give a number to describe your average pain over the last 24 hours.”* | Baseline, weeks 4, 8, 12, 16, 26, and 52 |
| Time to recovery | Pain diary (NPRS) (0-10, best to worst)<br>*“Record a number (0-10) to describe your average leg pain over the last 24 hours”* | Daily until recovery or up to 14 weeks |
| Quality of life | EQ-5D-5L [6] Visual Analogue Scale (VAS) (0-100, worst to best) | Baseline, weeks 4, 8, 12, 16, 26, and 52 |
|  | EQ-5D-5L Utility score (derived from a scoring algorithm with weights specific to Australian population [7]) (-0.301-1, worst to best) | Baseline, weeks 4, 8, 12, 16, 26, and 52 |
| Depression | Patient health Questionnaire – 9 [8, 9] (PHQ-9, 0-27, best to worst) | Baseline and week 8 |
| Anxiety | Generalised Anxiety Disorder 7-item [10] (GAD-7, 0-21, best to worst) | Baseline and week 8 |
| Sleep disturbance | Sleep Disturbance T-score (derived from the 8-item Patient-Reported Outcomes Information System (PROMIS) Sleep Disturbance 8a Short Form Adult v1.0) (see official document) | Baseline and week 8 |
| Participants' rating of global improvement | Global perceived effect (GPE) scale (-5 to 5, worst to best)<br>*“When you think about your leg pain, compared to before you started the study medication, how would you describe your leg pain these days?”* | Weeks 4, 8, 12, 16, 26, and 52 |
| Adverse events | Self-report | Weeks 4, 8, 12, 16 |
|  | Study pharmacist monitoring report | Week 2, 6, 12 |
|  | Doctor report | DREAM follow up GP survey |
|  | Additional AE Report | From randomisation to 1-year post-randomisation |
| Work absenteeism | Self-report | Baseline, weeks 4, 8, 12, 16, 26, and 52 |
| Adherence to study medication | Self-report and comparing the count of returned medications against the doctor's prescription. | Week 14 |
| Healthcare use and prescription medicines | Data linked from The Australian Medicare Benefits Schedule (MBS) and Pharmaceutical Benefits Scheme (PBS); self-report | Weeks 4, 8, 12, 16, 26, and 52 |
| ***Exploratory outcomes*** |  |  |
| Duloxetine serum trough concentration | Blood sample | Week 3 |
| Pharmacogenomic testing (eg, CYP2D6) | Buccal sample | Baseline |
| High-sensitivity c-reactive protein | Blood sample | Baseline, Week 3 |

### 4.7 Randomisation and blinding

Participants are randomised to receive either duloxetine or placebo at a 1:1 ratio using randomly permuted blocks of various sizes. Study medication packs are prepared according to the randomisation sequence by a clinical trials investigational product manufacturer (Syntro Health, Victoria, Australia), sealed, and distributed thereafter. The study medication packs are dispensed directly to the participant via a central pharmacy service provided by Syntro Health, except for those participants recruited from the hospital setting whose study medication is dispensed via the hospital’s clinical trials pharmacy. Participants are dispensed a study medication pack with a unique study enrolment number in sequential order. Randomisation occurs when the study medication pack is allocated to the participant after (i) the participant has met all inclusion criteria and none of the exclusion criteria, (ii) the informed consent process has been completed and consent has been obtained, and (iii) baseline data collection has been completed.

Allocation to group is concealed, and the active and placebo medicines are identical in appearance and taste, allowing blinding to the participant, treatment provider, assessor and all study personnel, including all Investigators and the Steering Committee. To test the success of blinding, participants are asked to guess their group allocation at the end of the dose maintenance phase at week 12, as either duloxetine, placebo or don’t know.

The statistical analysis will be performed blindly using randomly permuted (dummy) treatment allocations until all the data queries are resolved. The unblinded interim analysis will only remain available for the Data Safety Monitoring Board (DSMB) members until the formal unblinding.

### 4.8 Sample size

A sample size of 266 participants (133 per group) provides 90% power to detect a clinically worthwhile difference of 1 point (0-10) scale for leg pain intensity between the duloxetine and placebo groups. We assumed an SD of 2.5 based on previous duloxetine trials for back pain, and a two-tailed alpha of 5%. This sample size also provides >90% power to detect a clinically worthwhile difference of 3 points on the Roland-Morris Disability Sciatica Questionnaire, our key secondary outcome (assuming a conservative SD of 6).

To increase the chance of finding the correct answer early and minimise the expected sample size, we use an adaptive group sequential design with O’Brien-Fleming efficacy and non-binding futility stopping boundaries [11]. This approach is based on flexible ‘spending functions’ that adjust the level of alpha and beta spent based on the number of looks. It has the advantage that the exact number and timing of interim analyses do not need to be pre-specified. Based on 10,000 trial simulations with up to 3 optimally-spaced looks (2 interim analyses), this approach leads to an expected total sample size of 208 participants (an expected saving of 58 participants, or 22% of the sample size compared to a non-adaptive design), and in the case of no early stopping, a maximum sample size of 282 participants (only 16 [6%] more participants than with a non-adaptive design) (Figure 2). Allowing for a conservative estimate of 15% of dropouts, we will recruit a total sample size of 332 participants.

**Figure 2.**
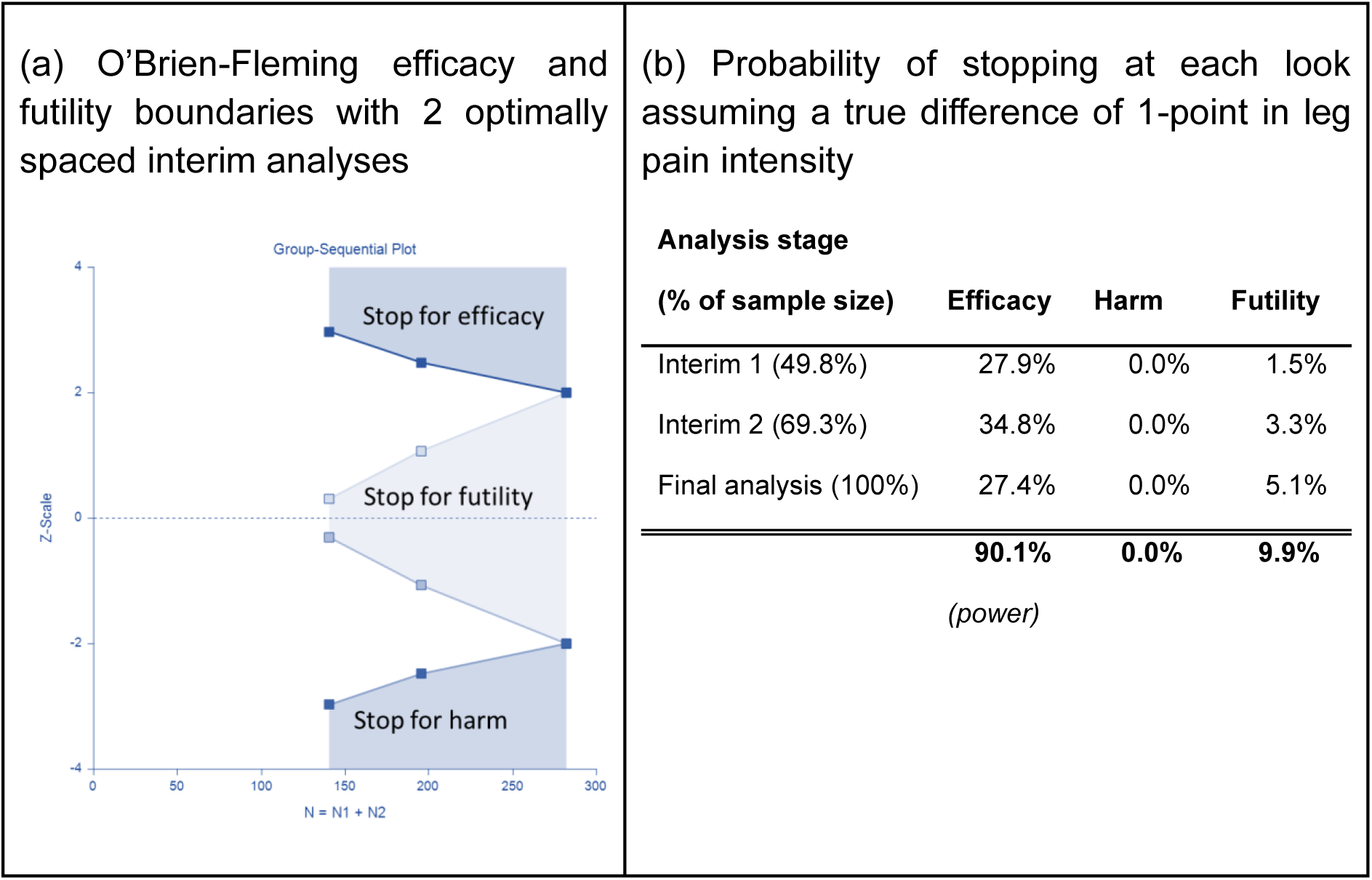
Giovanni.

## 5 Statistical analysis

### 5.1 Statistical hypotheses

The primary statistical hypotheses are as follows:

- **Null hypothesis**: The intervention, oral duloxetine, has the same effect as placebo in reducing leg pain in people with chronic sciatica.
- **Alternative hypothesis (2-sided)**: The intervention, oral duloxetine, is more effective than placebo in reducing leg pain in people with chronic sciatica.

### 5.2 Statistical principles

#### 5.2.1 Level of statistical significance

A group sequential design with three analyses is planned to be conducted when approximately 50%, 70%, and 100% of the total planned information has been accrued, although the actual timing may vary slightly (the number in parentheses indicate the approximate number of patients needed based on 15% with missing primary outcome data).

- 140 (50%) of participants have data for the primary time point at 12 weeks (approximately 166 patients recruited)
- 196 (70%) of participants have data for the primary time point at 12 weeks (approximately 230 patients recruited)
- 282 (100%) of participants have completed the study (approximately 332 patients recruited)

The design controls the overall two-sided type I error rate at 5% using an O’Brien–Fleming alpha-spending function [11]. It is powered to achieve approximately 90% power to detect the pre-specified treatment effect by the final analysis under the alternative hypothesis.

At each analysis, the primary hypothesis will be tested using the corresponding significance level derived from the alpha and beta spending function. Early termination for efficacy or futility will be recommended if the boundary is crossed. The decision of efficacy or futility will be based on the z-score thresholds and corresponding p-values (see Table 3), while all the confidence intervals will still be shown as 95%. At each interim analysis, the boundary thresholds and corresponding p-values should be updated with the actual sample size.

**Table 3.** Stopping rules.

| Analysis stage | Interim |  | Final |
| --- | --- | --- | --- |
| Planned information rate | 50%<br>(N = 140) | 70%<br>(N = 196) | 100%<br>(N = 282) |
| Cumulative alpha spent | 0.3% | 1.4% | 5.0% |
| Cumulative beta spent | 1.1% | 3.7% | 10.0% |
| Cumulative power | 27.9% | 62.7% | 90.1% |
| Significance threshold (two-sided) | 0.3% | 1.4% | 4.6% |
| Efficacy boundary (z-value scale) | 2.969 | 2.477 | 2.000 |
| Futility boundary (z-value scale) | 0.307 | 1.067 |  |

For the final analysis, analyses of the primary outcome (leg pain intensity at week 12) will be unadjusted for test multiplicity; however, it will still be adjusted for the potential alpha spent at the interim analyses i.e. if interim analyses were conducted and the study did not stop early. Therefore, the intervention effect will be considered statistically significant at the final analysis if the two-sided p-value is less than 0.046, reflecting the adjustment for the planned interim analyses. For secondary outcomes (one family), the family-wise error rate will be controlled using a Holm-Bonferroni correction [12]. No other multiplicity adjustment will be applied. Exploratory outcomes will be reported descriptively, and no adjustment will be necessary.

#### 5.2.2 Interim analysis principles

Two interim analyses are planned when approximately 50% and 70% of the total of participants have data for primary outcome at 12-weeks. Actual timing may vary slightly due to the pace of recruitment and data availability. Interim analyses are intended solely to inform the independent DSMB regarding potential early stopping for efficacy, futility, or harm (see Figure 2). As a result of ongoing data collection and incomplete follow-up at interim time points, the statistical methods used at interim analyses will differ from those applied in the final analysis.

##### Primary outcome analysis at interim

For the primary outcome, both the base model (section 5.6.1) and the adjusted model (section 5.6.2) will be fitted at each interim analysis.

Interim analyses will use the following approach:

- Analyses will be performed using the ITT population only (section 5.3).
- No imputation will be performed.
- No sensitivity analyses (section 5.6.4) and no subgroup analyses (section 5.6.5) will be conducted.

##### Secondary outcomes at interim

Baseline characteristics, follow-up information, and all secondary outcomes will be summarised descriptively at each interim analysis. No hypothesis testing or statistical modelling of secondary outcomes is planned at interim time points. These summaries are provided exclusively to support DSMB review of safety and trial conduct.

**Table 4.** Comparison of interim vs final analyses.

| Aspect | Interim analyses | Final analysis |
| --- | --- | --- |
| Timing Conducted | when ~50% and ~70% of total planned information is accrued (actual timing may vary) | Conducted after full data collection (or after all currently enrolled participants complete follow up if early stopping occurs) |
| Purpose | To inform DSMB decisions regarding early stopping for efficacy, futility, or safety | To generate definitive estimates and inferences addressing primary and secondary study objectives |
| Analysis population | ITT population only | Full ITT population; per protocol or other analysis sets may also be used for sensitivity analyses |
| Primary outcome model | Base and adjusted models | Adjusted model (primary), with additional models as sensitivity analyses |
| Handling of missing primary outcome data | No imputation | Multiple imputation, if required |
| Sensitivity analyses | Not planned | Full suite of sensitivity analyses performed as prespecified (Section 5.6.4 ) |
| Subgroup analyses | Not planned | All prespecified subgroup analyses performed (Section 5.6.5 ) |
| Secondary outcomes | Reported descriptively only; no modelling or formal inference | Analysed using prespecified methods (observed data only; exploratory) |
| Use of results | Contributes to group sequential stopping decisions while maintaining Type I error control | Used to estimate treatment effects, perform hypothesis testing, and draw final conclusions |
| Reporting | Provided to DSMB only; results remain blinded to investigators and trial team | Full statistical reporting in final study outputs for publications |

##### Transition to final analysis

If the trial is stopped early based on interim findings, the final analysis may proceed once all currently enrolled participants have completed their scheduled follow-up, ensuring complete data collection for the cohort already randomised at the time of stopping.

The final analysis will then be performed using the full pre-specified statistical methods, including multiple imputation (as applicable), sensitivity analyses, and subgroup analyses.

#### 5.2.3 Statistical software

Analyses will be conducted primarily using SAS (version 9.3 or above) or R (version R 4.3.1 or above).

### 5.3 Analysis populations

Intention-to-treat (ITT) analysis set: All randomised participants will be analysed according to their randomised group, regardless of adherence to the protocol, and excluding those who withdrew their consent. This will be the primary analysis set to assess efficacy and safety in both interim and final analyses.

Per Protocol (PP) analysis set: All participants in the ITT dataset without any intercurrent events related to adherence to study treatment (see Table 5). The PP set will be used in only final analysis to re-run analyses of the primary outcome, including sensitivity analyses, as well as the analysis of SAEs.

**Table 5.**
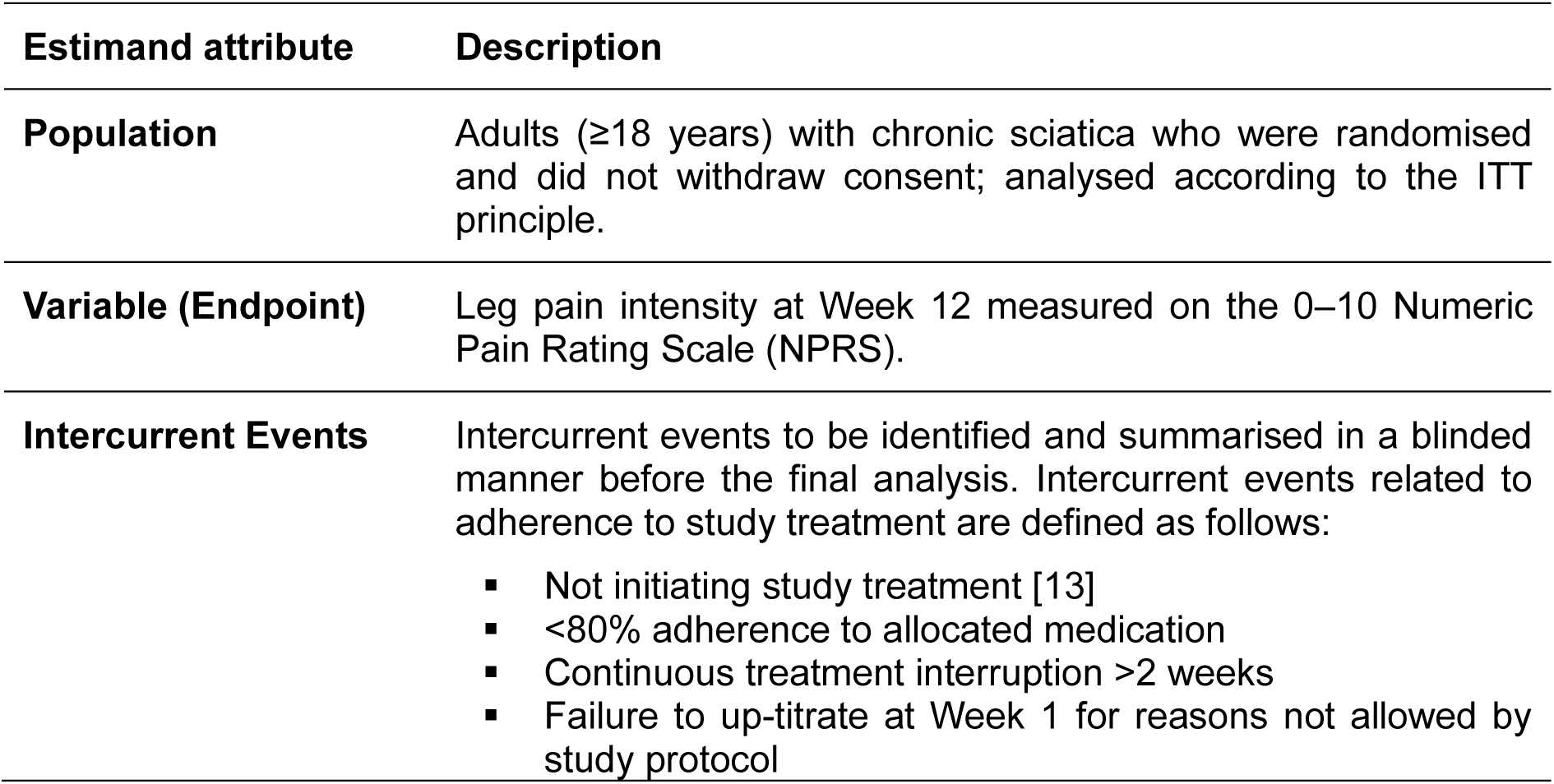
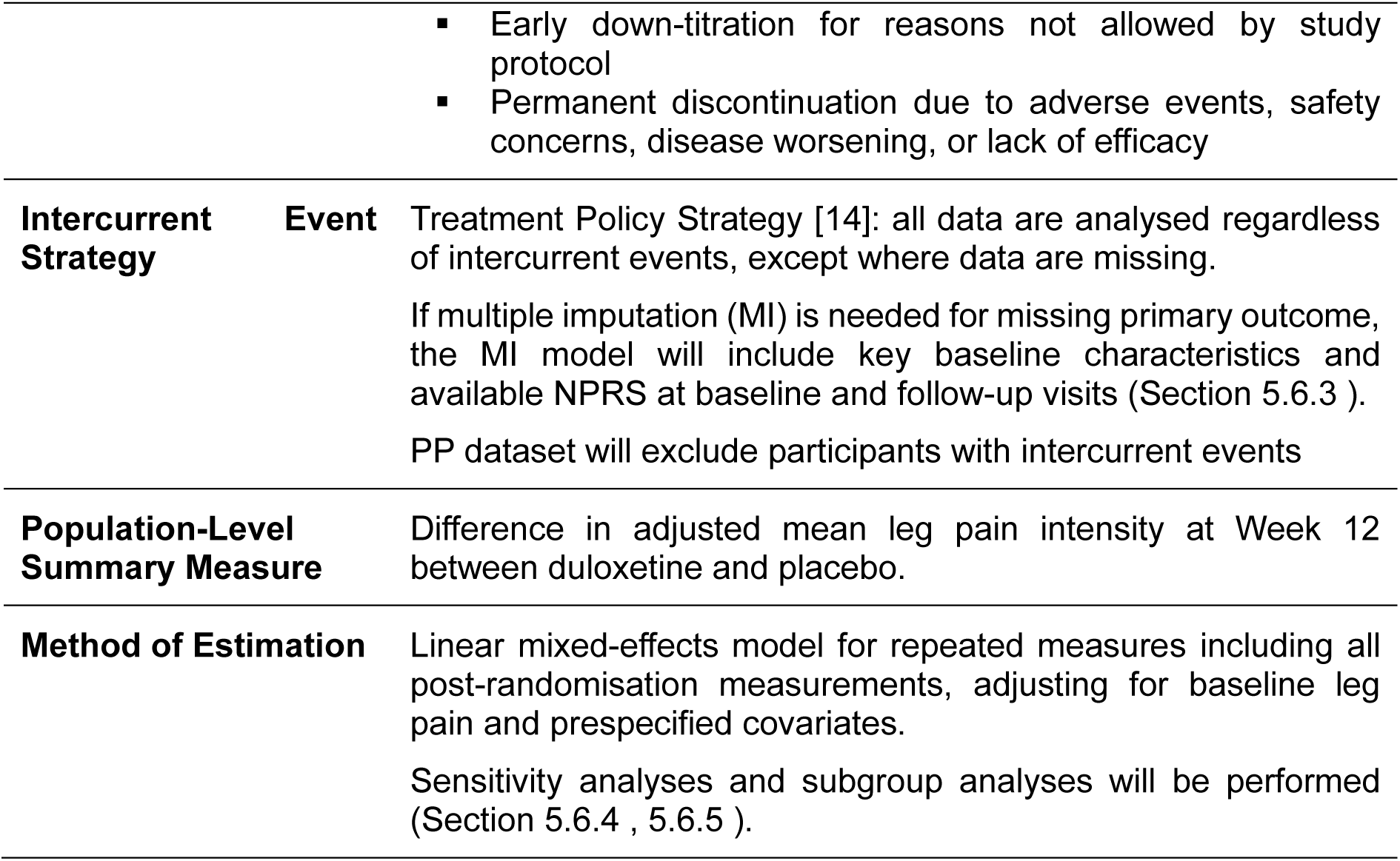
Primary estimand.

### 5.4 Data description

Baseline characteristics and all the outcome variables will be presented by randomisation group. Discrete variables will be summarised by frequencies and percentages. Percentages will be calculated for participants whose data are available. Continuous variables will be summarised by using mean and SD, and median and interquartile range (Q1-Q3).

### 5.5 Estimands

#### 5.5.1 Primary estimand

The primary estimand is summarised in Table 5.

#### 5.5.2 Secondary estimands

For the analyses of secondary outcomes (Section 5.7), a Treatment Policy Strategy will be applied [14].

Analyses will be conducted using observed data only and will be performed during the final analysis. Intercurrent events will not be explicitly accounted for beyond their contribution to any missing data. Results will be interpreted with caution and used to support understanding of the broader impact of duloxetine on pain, disability, and quality of life.

### 5.6 Analysis of the primary outcome

For the final analysis, the primary outcome will be analysed in both the ITT and PP sets.

For the interim analyses, the primary outcome will be analysed in the ITT set only. We will conduct the analysis after we have 50% of participants (or 70% for the second interim) followed up at week 12.

#### 5.6.1 Base model

The primary outcome will be analysed using a repeated-measure linear mixed model including leg pain intensity scores at each post-randomisation follow-up until week 12, which allows every participant with at least one measurement to be included. This model will include the randomised intervention, the baseline leg pain intensity score, and the follow-up time points as categorical variables. An interaction term between time point and intervention will be included to allow the intervention effect to be estimated separately at each time point. The effect of the intervention at 12 weeks post-randomisation (primary time point) will be estimated from the model as the mean difference (MD) and its 95% confidence interval (95% CI).

#### 5.6.2 Adjusted model

An adjusted model will be built by adding age, sex, time from sciatica onset to randomisation and depression (PHQ9) to the base model.

The adjusted model will be the primary analysis for the primary outcome (Table 6).

**Table 6.**
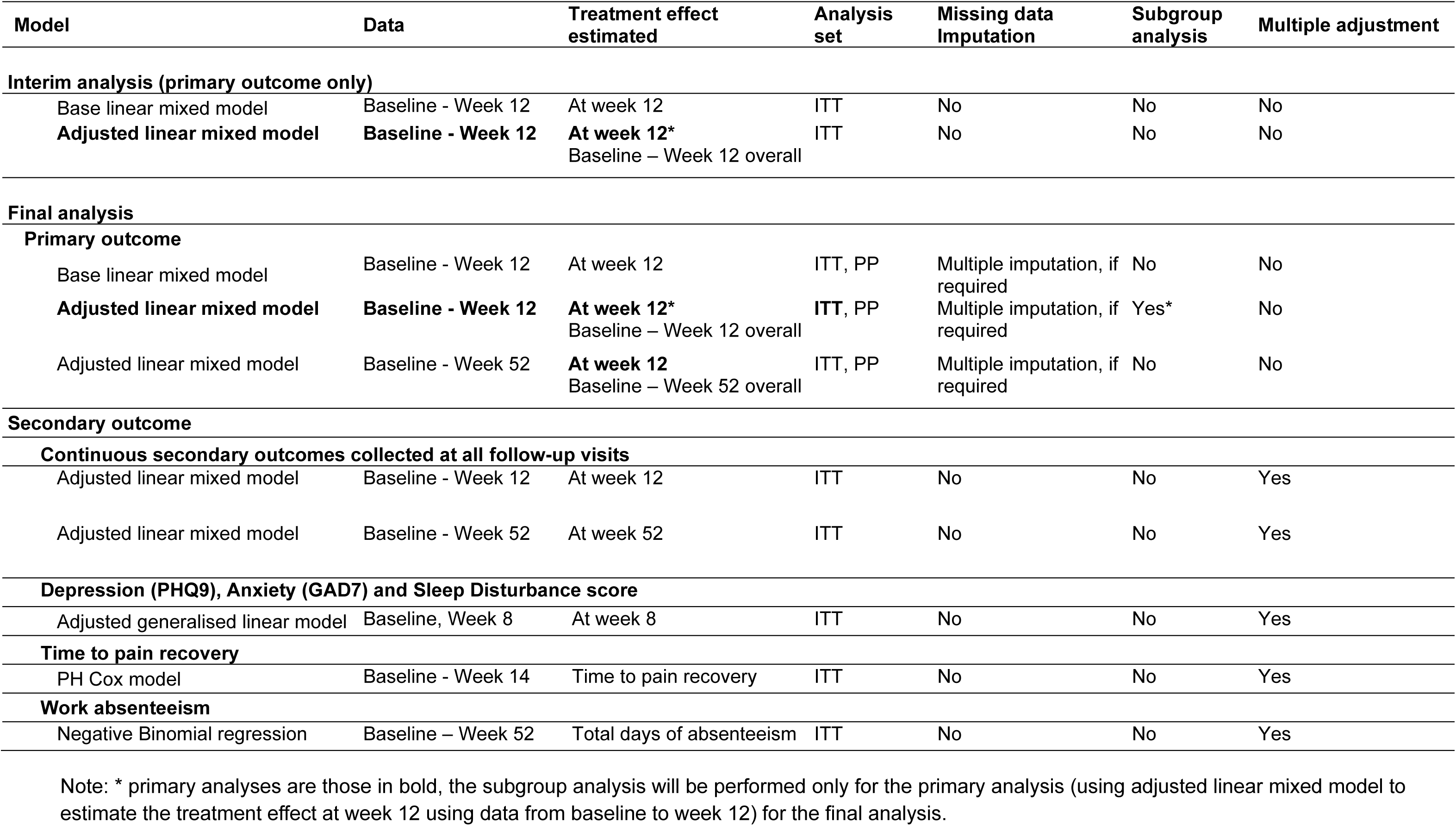
Summary of the analytical methods.

| Model | Data | Treatment effect estimated | Analysis set | Missing data imputation | Subgroup analysis | Multiple adjustment |
| --- | --- | --- | --- | --- | --- | --- |
| **Interim analysis (primary outcome only)** |  |  |  |  |  |  |
| Base linear mixed model | Baseline - Week 12 | At week 12 | ITT | No | No | No |
| **Adjusted linear mixed model** | **Baseline - Week 12** | **At week 12\***<br>Baseline – Week 12 overall | ITT | No | No | No |
| **Final analysis** |  |  |  |  |  |  |
| **Primary outcome** |  |  |  |  |  |  |
| Base linear mixed model | Baseline - Week 12 | At week 12 | ITT, PP | Multiple imputation, if required | No | No |
| **Adjusted linear mixed model** | **Baseline - Week 12** | **At week 12\***<br>Baseline – Week 12 overall | **ITT, PP** | Multiple imputation, if required | Yes* | No |
| Adjusted linear mixed model | Baseline - Week 52 | **At week 12**<br>Baseline – Week 52 overall | ITT, PP | Multiple imputation, if required | No | No |
| **Secondary outcome** |  |  |  |  |  |  |
| **Continuous secondary outcomes collected at all follow-up visits** |  |  |  |  |  |  |
| Adjusted linear mixed model | Baseline - Week 12 | At week 12 | ITT | No | No | Yes |
| Adjusted linear mixed model | Baseline - Week 52 | At week 52 | ITT | No | No | Yes |
| **Depression (PHQ9), Anxiety (GAD7) and Sleep Disturbance score** |  |  |  |  |  |  |
| Adjusted generalised linear model | Baseline, Week 8 | At week 8 | ITT | No | No | Yes |
| **Time to pain recovery** |  |  |  |  |  |  |
| PH Cox model | Baseline - Week 14 | Time to pain recovery | ITT | No | No | Yes |
| **Work absenteeism** |  |  |  |  |  |  |
| Negative Binomial regression | Baseline – Week 52 | Total days of absenteeism | ITT | No | No | Yes |

Note: \* primary analyses are those in bold, the subgroup analysis will be performed only for the primary analysis (using adjusted linear mixed model to estimate the treatment effect at week 12 using data from baseline to week 12) for the final analysis.

#### 5.6.3 Missing data handling

Missing data will be examined for the analysis of the primary outcome. We will report the percentage of participants missing NPRS at baseline and each follow-up visit. We will also report the number of participants included in the primary analysis, i.e., having all the covariates (e.g., age, sex) and at least one of the NPRS at baseline and follow-up visits.

The primary analysis will include all participants with at least one valid NPRS measurement post-randomisation and with no missing baseline covariate. This approach will provide valid inference under the missing at random (MAR) assumption.

If the proportion of participants excluded from the primary analysis is ≤ 5% of all the randomised participants, no imputation will be performed. Otherwise, we will perform multiple imputation (MI) assuming missing at random (MAR).

The MI model will include the treatment arm, key baseline characteristics (age, gender, BMI, smoking, time from sciatica onset to randomisation (month), leg pain intensity, depression, anxiety, sleep disturbance), and available NPRS at baseline and follow-up visits. One hundred sets will be imputed using a fully conditional specification with discrete variables imputed using a discriminant function [15]. The imputed data will be analysed using the main analysis, i.e. the adjusted model.

For the interim analysis, no imputation is planned.

#### 5.6.4 Sensitivity analyses

For the final analysis, if the missing data from the primary analysis is ≤ 5%, the adjusted model results using available data will remain the primary analysis. If the missing data is greater than 5%, the adjusted model results using imputed data will be considered as the primary analysis, while the one using available data will be a sensitivity analysis.

The primary analysis, i.e., the adjusted linear mixed model, will also produce the overall treatment effect throughout the follow-up period until week 12, which will be reported as a sensitivity analysis. For the final analysis, we will also apply the same adjusted model using data from baseline to week 52 to estimate the treatment effect at week 12, as well as the overall treatment effect from baseline to week 52.

#### 5.6.5 Subgroup analyses

Potential interactions with NPRS will be explored using the following baseline variables. The subgroup analyses are exploratory and will be carried out irrespective of whether there is a statistically significant treatment effect on the primary outcome.

- Age groups (< 65 years / ≥ 65 years)
- Sex (Female / Male)
- Depressive symptoms (PHQ-9 <10 / ≥ 10) [9]
- Anxiety symptoms (GAD-7 <10 / ≥ 10) [16]
- Sleep disturbance (PROMIS short-form V1.0 Sleep Disturbance 8a <60 / ≥60) (see <u>documentation</u>)

The analysis for each subgroup will be performed by adding the subgroup variable, as well as its interaction with the intervention as a fixed effect to the main analysis, i.e., adjusted mixed model in Section 5.6.2. Collinearity will be avoided when adding the subgroup variable, e.g., when age group is added, the age as a continuous variable in the main model will be removed. Within each subgroup, summary measures will include raw numbers and percentages of the participants within each treatment arm, as well as the MD for treatment effect with a 95% CI. The results will be displayed in a forest plot including the p-values for heterogeneity corresponding to the interaction term between the intervention and the subgroup variables.

Since these subgroup analyses are exploratory, they will only be performed in the final analysis on the non-imputed data (i.e. using all data available).

### 5.7 Analysis of the secondary outcomes

The statistical modelling of the secondary outcomes is not needed for interim analysis and will be performed only for the final analysis, in the ITT set only (see Table 6).

#### 5.7.1 Continuous outcomes

For the continuous outcomes collected at baseline, 4, 8, 12, and 16 weeks, including disability, quality of life, low back pain intensity, and GPE; similarly to the primary analysis, a linear mixed model with adjustment for outcome variable at baseline, age, sex, sciatica duration will be used. The effect of the intervention at 12 weeks post-randomisation (primary time point) will be estimated from the model as the MD and its 95% CI.

We will apply the same adjusted model using data from baseline to week 52 to estimate the treatment effect at week 52 (see Table 6). For PHQ9, GAD7 and PROMIS Sleep Disturbance T-score, continuous outcomes collected at baseline and 8 weeks, generalised linear models will be used, including the randomised intervention and adjusting for age, sex and the baseline value of the outcome.

#### 5.7.2 Time-to-event outcomes

Participants are asked to record the average daily leg pain intensity score in a pain diary for the duration of treatment (14 weeks) using a 0-10 NPRS. Recovery is defined as 7 consecutive days with leg pain intensity ≤1 out of 10. The day of ‘event’ will be the first day of recovery, i.e. day 1 of the seven consecutive days. Data are censored at 14 weeks or if participants report having recovered, whichever occurs first (which means that if a pain recovery is defined, the first day with leg pain intensity ≤1 should occur before or on day 98).

Medians and quartiles of time to recovery will be obtained from the cumulative incidence functions. The effect of the intervention will be estimated as the hazard ratio (HR) and its 95% CI using a proportional Cox model, which includes a fixed effect of the group allocation, baseline leg pain intensity, sex, age, sciatica duration, and depression symptoms.

No imputation, covariate adjustment, subgroup analyses or other sensitivity analyses will be applied to this outcome.

#### 5.7.3 Count outcomes

At weeks 4, 8, 12, 16, 26, and 52, participants are asked whether they took time off work because of their sciatica since their last follow-up. At weeks 4, 8, 12, and 16, participants are asked to recall the previous four weeks. At week 26, the previous 10 weeks (since week 16). At week 52, the recall period is 6 months (since week 26). Participants replying yes are prompted to estimate the number of absent days. The number of absent days from baseline to week 52 will be summed.

The effect of the intervention on work absenteeism will be analysed using a negative binomial (NB) regression model. Work absenteeism will be defined as the number of days off work in the previous four weeks because of sciatica. The intervention effect will be expressed as a rate ratio (RR) with a 95% confidence interval.

The model will include treatment group and will be adjusted for age, sex, time from sciatica onset to randomisation, depression measured using the PHQ-9, and baseline absenteeism. The NB model is preferred over a Poisson model because absenteeism data are expected to be overdispersed, with the variance exceeding the mean. This analysis will be limited to participants who were working at baseline; participants who were not employed at baseline will be excluded.

#### 5.7.4 Adverse events

Adverse events (AEs) are defined as any untoward medical occurrence that does not necessarily have a causal relationship with this treatment. Serious adverse events (SAEs) are any untoward medical occurrence that: (I) results in death, (ii) is life-threatening, (iii) requires hospitalisation or prolongation of existing hospitalisation, (iv) results in persistent or significant disability or incapacity, (v) is a congenital anomaly or birth defect, or is a medically significant or important event or reaction. Serious adverse events are assessed for relatedness (yes/no) and expectedness (yes/no) by an independent medical monitor.

Participants are asked to report any AE or SAE at weeks 2, 4, 6, 8, 12, and 16. At weeks 2, 6, and 12, participants complete the validated Antidepressant Side Effect Checklist [17] which includes 21 common adverse reactions to antidepressants. At weeks 4, 8, 12, and 16, participants complete an open-ended question to collect adverse event data. Adverse events can also be reported to study doctors at follow-up visits. Where study doctors and participants have reported the same adverse event on the same date, we will exclude the doctor data to avoid duplication. If participants report adverse events directly to research staff at any time via other means (e.g., text messages), these data are recorded in an additional form in the trial database.

We will use the Medical Dictionary for Regulatory Activities (MedDRA) to code all AEs and SAEs. Verbatim terms will be coded to the most appropriate Lowest Level Term (LLT) and linked to a corresponding Preferred Term (PT), which will be used for analysis and reporting. We will also use System Organ Classes (SOC) for summarising safety data.

The proportion of participants with adverse events and serious adverse events overall will be compared between groups using the Chi-square test.

### 5.8 Cost-effectiveness

#### 5.8.1 Overview

The economic evaluation will determine the cost-effectiveness and cost-utility of duloxetine compared with placebo in people with sciatica. The primary objective of the economic evaluation is to determine the cost-utility at 12 months using the EQ-5D-5L and a published value set [7]. The secondary objective is to estimate the within-trial cost effectiveness at 12 months using the primary outcome. The economic evaluation will be planned, conducted, and reported in accordance with the Consolidated Health Economic Evaluation Reporting Standards (CHEERS) [18].

The within-trial economic analysis will be performed using individual patient level data from the DREAM trial and linked Medical- and Pharmaceutical- Benefits Scheme data. The analytical approaches will take the form of cost-effectiveness and cost-utility analysis. Incremental cost-effectiveness and cost-utility ratios will be calculated by taking a ratio of the difference in the mean costs and mean effects (or utility measure). The base case analyses will take an Australian Health System perspective.

#### 5.8.2 Economic data collection

Resource use will be collected from multiple sources. Administratively linked data will be used for Medicare: health services via the Medical Benefits Scheme (MBS) and prescription medications via the Pharmaceutical Benefits Scheme (PBS). Patient diaries will identify the non-PBS medications (private scripts) and over-the-counter (OTC) medications as these can be significant components of pain management not captured in the PBS data. Hospitalisations will also be self-reported.

MBS and PBS data from randomisation to 52 weeks post-randomisation will be obtained at the end of trial for consenting participants. Self-reported medications and health service use in the last 4 weeks will be recorded every four weeks from baseline to week 16, and at 26-, and 52-weeks using trial case report forms.

MBS and PBS costs will be determined from the relevant item number and current schedule at the time of analysis. OTC medications will be costed based on pharmacist websites (e.g. Chemist Warehouse) and hospitalisations using the Independent Health and Aged Care Pricing Authority (IHACPA) cost weights from the National Hospital Cost Data Collection.

The primary economic outcome measure will be Quality-Adjusted Life Years (QALYs). These will be derived from participant responses to the EQ-5D-5L quality of life instrument (collected at baseline, 4, 8, 12, 16, 26, and 52 weeks) and the Australian utility value set [7]. The responses will be transformed into QALYs using the area-under-the-curve method adjusting for baseline differences [19, 20]. Missing data will be handled as described in section 5.6.3 in the first instance, noting that exploration of the handling of missing data is part of sensitivity analyses described in the next section.

#### 5.8.3 Economic data analysis

The analysis population for the base-case analysis will be all randomised participants as per the ‘intention to treat’ principle, with a ‘per protocol’ set for sensitivity analysis. The analysis will take place post-trial once the MBS and PBS data are available, there will be no interim economic data analysis. No discounting will be applied as the time-horizon is 12 months. The cost-utility analysis will use a threshold of $50,000 AUD per QALY reflecting the unofficial Australian threshold [21].

Mean differences in costs, QALYs and net benefits between the treatment groups will be estimated with associated 95% confidence intervals. There is limited guidance on health economic analysis alongside trials with group sequential design, and the bias which may result from this design. We will follow existing guidance to generate adjusted estimates and confidence intervals [22]. Should further guidance become available we will endeavour to incorporate these.

Differences in health service use will be described but not statistically compared.

Adjusted mean difference in QALYs and adjusted difference in overall mean costs between the arms will be analysed using Bias Adjusted Maximum Likelihood Estimate as described by Flight [22]. Cost and QALY data will be combined to calculate an incremental cost-effectiveness ratio (ICER). The nonparametric bootstrapping approach will be used to determine the level of sampling uncertainty surrounding the mean ICER by generating 10,000 estimates of incremental costs and benefits.

Face validity tests will be conducted on data (e.g. to identify misspelt text) and checked with study staff. Corrections made will be documented in the statistics program code.

Flexible Bayesian longitudinal models, as described by Mason et al. [23], will be used in sensitivity analysis to consider the impact of informative missing data (such as that from group sequential design) on the interpretation and conclusions of the cost-effectiveness analysis. Analyses will also be conducted on the final dataset to investigate how cost-effectiveness varies between different patient subgroups (as described in section 5.6.5). Any subgroup analyses for which the smaller subgroup includes fewer than 50 participants will be omitted.

### 5.9 Sub-studies

Several sub-studies will be conducted using trial data. These include (i) a causal mediation analysis to investigate potential mechanisms of duloxetine [24], (ii) a complier average causal effect (CACE) analysis to investigate treatment efficacy in participants who complied with the treatment regimen [25], (iii) an exploratory study investigating the moderating effect of pharmacogenes and high-sensitive c-reactive protein (hs-CRP) on trial outcomes in a subset of participants, and (iv) an exploratory study investigating the influence of duloxetine serum trough concentration on clinical outcomes in a subset of participants. We will prepare analysis plans for these studies prior to initiating analyses and will report findings in detail in later publications.

## Data Availability

No data were generated or analysed as part of this statistical analysis plan. Data availability will be described in the final study report and related publications, in accordance with applicable data-sharing policies and participant consent.

## 5 List of abbreviations

ABBREVIATION

95% CI: 95% Confidence Interval
AE: Adverse event
DSMB: Data Safety Monitoring Board
GAD-7: Generalised Anxiety Disorder 7-item
GPE: Global Perceived Effect
hsCRP: high-sensitivity C-reactive protein
IMP: Investigational Medicinal Product
MD: Mean Difference
MBS: Medicare Benefits Schedule
MI: Multiple imputation
NB: Negative Binomial
NPRS: Numerical Pain Rating Scale
PBS: Pharmaceutical Benefits Scheme
PHQ-9: Patient Health Questionnaire-9
PROMIS: Patient-Reported Outcomes Measurement Information System
RMDQ: Roland Morris Disability Questionnaire
SAE: Serious Adverse Event
SD: Standard deviation
DREAM - Statistical Analysis Plan: Version 1.1 – July 2026

## Notes

### Competing Interest Statement

The study has been awarded funding from the NHMRC, Australia. The investigators maintain full autonomy in the design, conduct and reporting of the study. We have ethics approval to reimburse study clinicians and participants for their time spent on study- specific tasks. NBF has received consultancy fees from NeuroPN, Saniona, PharmNovo, and Tris Pharma, and has undertaken consultancy work for Aarhus University with remunerated work for AKIGAI, Merz, and NovoNordisk. MU is the chief investigator or co- investigator on multiple previous and current research grants from the UK National Institute for Health Research and is a co- investigator on grants funded by the Australian NHMRC and Norwegian MRC. He is a director and shareholder of Clinvivo Ltd. that provides electronic data collection for health services research. He receives some salary support from University Hospitals Coventry and Warwickshire. He is a co- investigator on two current and one completed NIHR funded studies that have, or have had, additional support from Stryker Ltd. He has accepted travel expenses for speaking at academic meetings.

### Clinical Trial

ACTRN12624000919516

### Funding

- National Health and Medical Research Council: APP2023931

## References

1. McLachlan, H., et al., DREAM: an adaptive, randomised, placebo-controlled trial of duloxetine for reducing leg pain in people with chronic sciatica-trial protocol. BMJ Open, 2024. 14(12): p. e096796.

2. Ivanova, J.I., et al., Real-world practice patterns, health-care utilization, and costs in patients with low back pain: the long road to guideline-concordant care. The Spine Journal, 2011. 11(7): p. 622–632.

3. Ferreira, G.E., et al., Patterns of antidepressant use in people with low back pain: A retrospective study using workers’ compensation data. Eur J Pain, 2025. 29(1): p. e4773.

4. Ferreira, G.E., et al., Efficacy and safety of antidepressants for the treatment of back pain and osteoarthritis: systematic review and meta-analysis. Bmj, 2021. 372: p. m4825.

5. Patrick, D.L., et al., Assessing health-related quality of life in patients with sciatica. Spine, 1995. 20(17): p. 1899–1908.

6. Herdman, M., et al., Development and preliminary testing of the new five-level version of EQ-5D (EQ-5D-5L). Quality of life research, 2011. 20(10): p. 1727–1736.

7. Norman, R., et al., The use of a discrete choice experiment including both duration and dead for the development of an EQ-5D-5L value set for Australia. Pharmacoeconomics, 2023. 41(4): p. 427.

8. Kroenke, K., R.L. Spitzer, and J.B. Williams, The PHQ-9: validity of a brief depression severity measure. Journal of general internal medicine, 2001. 16(9): p. 606–613.

9. Negeri, Z.F., et al., Accuracy of the Patient Health Questionnaire-9 for screening to detect major depression: updated systematic review and individual participant data meta-analysis. bmj, 2021. 375.

10. Spitzer, R.L., et al., A brief measure for assessing generalized anxiety disorder: the GAD-7. Archives of internal medicine, 2006. 166(10): p. 1092–1097.

11. O’Brien, P.C. and T.R. Fleming, A multiple testing procedure for clinical trials. Biometrics, 1979. 35(3): p. 549–56.

12. Holm, S., A Simple Sequentially Rejective Multiple Test Procedure. Scandinavian Journal of Statistics, 1979. 6(2): p. 6.

13. Kahan, B.C., et al., Using modified intention-to-treat as a principal stratum estimator for failure to initiate treatment. Clinical Trials, 2023. 20(3): p. 269–275.

14. Clark, T.P., et al., Estimands: bringing clarity and focus to research questions in clinical trials. BMJ open, 2022. 12(1): p. e052953.

15. van Buuren, S., Multiple imputation of discrete and continuous data by fully conditional specification. Stat Methods Med Res, 2007. 16(3): p. 219–42.

16. Akturk, Z., et al., Generalized Anxiety Disorder 7-item (GAD-7) and 2-item (GAD-2) scales for detecting anxiety disorders in adults. Cochrane Database Syst Rev, 2025. 3(3): p. CD015455.

17. Uher, R., et al., Adverse reactions to antidepressants. Br J Psychiatry, 2009. 195(3): p. 202–10.

18. Husereau, D., et al., Consolidated Health Economic Evaluation Reporting Standards 2022 (CHEERS 2022) statement: updated reporting guidance for health economic evaluations. MDM Policy & Practice, 2022. 7(1): p. 23814683211061097.

19. Drummond, M.F., et al., Methods for the economic evaluation of health care programmes. 2015: Oxford university press.

20. Manca, A., N. Hawkins, and M.J. Sculpher, Estimating mean QALYs in trial-based cost-effectiveness analysis: the importance of controlling for baseline utility. Health economics, 2005. 14(5): p. 487–496.

21. Taylor, C. and S. Jan, Economic evaluation of medicines. Australian prescriber, 2017. 40(2): p. 76.

22. Flight, L., The use of health economics in the design and analysis of adaptive clinical trials. 2020, University of Sheffield.

23. Mason, A.J., et al., Flexible Bayesian longitudinal models for cost-effectiveness analyses with informative missing data. Health economics, 2021. 30(12): p. 3138–3158.

24. Cashin, A.G., et al., *Understanding how health interventions or exposures produce their effects using mediation analysis*. bmj, 2023. 382.

25. Yang, Z., et al., Oxycodone/naloxone versus placebo for acute spinal pain in compliant participants: secondary analysis of a randomised clinical trial. British Journal of Anaesthesia, 2026.

